# *CREBBP* variants underlying Menke–Hennekam syndrome exhibit histone acetyltransferase hyperactivity with divergent phase behaviors

**DOI:** 10.64898/2026.10.26.26365886

**Authors:** Taiga Aoki, Yasuyuki Fukuhara, Tetsumin So, Arisa Igarashi, Masahiko Yamamori, Kumiko Yanagi, Tomoko Kawai, Rika Kosaki, Kenji Kurosawa, Yoichi Matsubara, Tadashi Kaname

**Affiliations:** Department of Genome Medicine, National Center for Child Health and Development, Tokyo, Japan; Center for Maternal-Fetal, Neonatal and Reproductive Medicine, National Center for Child Health and Development, Tokyo, Japan; Advanced Pediatric Medicine, Tohoku University School of Medicine, Sendai, Japan; Division of Medical Genetics, National Center for Child Health and Development, Tokyo, Japan; Department of Maternal-Fetal Biology, National Research Institute for Child Health and Development, Tokyo, Japan; CRIFM Prenatal Medical Clinic; Institute of Resource Development and Analysis, Kumamoto University, Kumamoto, Japan

**Keywords:** rare disease, histone acetyltransferase, liquid–liquid phase separation, transcriptional condensates

## Abstract

Menke–Hennekam syndrome (MKHK) is a neurodevelopmental disease characterized by specific pathogenic regions within the transcriptional coactivator *CREB-binding protein* (*CREBBP*). CREBBP regulates gene expression through histone acetyltransferase (HAT) activity and involves IDR-dependent transcriptional condensates. Although domain-specific phenotypes and distinct DNA methylation episignatures have been recognized in MKHK, the underlying molecular mechanisms remain widely unclear. In this study, we identified a recurrent missense variant, p.(R1868W) [R1868W] and a novel splice variant r.5963_5969del,p.(Cys1723Thrfs*19) [C1723fs] in two families clinically diagnosed with MKHK and manifesting developmental delay, characteristic facial features, and growth impairment. R1868W and C1723fs exhibited increased auto-, nonhistone protein, and histone acetylation. Despite these shared enzymatic abnormalities, the variants promoted divergent disruption in liquid–liquid phase separation. The recurrent CBP-ID4 variant R1868W activated the formation of low-mobility aberrant nuclear condensates, whereas the C-terminal, truncating variant C1723fs, impaired condensate formation and showed diffuse nuclear localization. Treatment with the selective p300/CBP HAT inhibitor A485 partially restored condensate dynamics and fusion-relaxation behavior in R1868W. Furthermore, R1868W recruited BRD4 around condensates in an acetylation-dependent manner, which was suppressed by the BET inhibitor JQ1. Together, these findings suggest that aberrant condensate material properties arising from disrupted CREBBP IDR-dependent regulation play a critical role in the molecular pathogenesis of MKHK.

**Significance Statement:** Pathogenic variants in specific regions of the transcriptional coactivator CREBBP underlie the Menke–Hennekam syndrome (MKHK), but its molecular basis is poorly understood. Here, we show that two *CREBBP* variants responsible for MKHK share a common feature of increased histone acetyltransferase (HAT) activity but disrupt liquid–liquid phase separation in distinct ways. The recurrent CBP-ID4 missense variant R1868W promoted the formation of low-mobility aberrant condensates and acetylation-dependent recruitment of BRD4, as opposed to the C-terminal truncating variant C1723fs. Notably, HAT inhibition restored condensate dynamics in R1868W, suggesting that condensate properties are reversibly regulated by enzymatic activity. These findings reveal a framework linking pathogenic variants affecting CREBBP IDR-dependent regulation to aberrant condensate material properties in MKHK.

## Introduction

Intracellular membraneless organelles spatially participate in biochemical reactions, such as transcription and RNA processing. Many of these structures are organized during liquid–liquid phase separation (LLPS), in which proteins and nucleic acids spontaneously condense into concentrated phases (1–4).

LLPS is governed by the balance between intermolecular interactions that facilitate condensation (enthalpy) and entropy that favors homogeneous mixing. In proteins, multivalency arising from multiple weak interactions plays a central role in driving phase separation such that subtle alterations in intermolecular interactions can markedly affect phase behavior. Intrinsically disordered regions (IDRs), which lack stable three-dimensional structures, are particularly important for LLPS given that they mediate dynamic and multivalent interactions (5,6). Recent studies have demonstrated that amino acid sequence patterns (i.e., “sequence grammar” of IDRs) and post-translational modifications within IDRs regulate the properties and subcellular localization of phase-separated condensates. Various features, such as charge distribution, polyglutamine tract (polyQ), and aromatic residue-mediated interactions, influence the strength and selectivity of intermolecular interactions, thereby determining phase behavior (7–9). Conversely, alterations in these sequence features can induce aberrant condensate formation or impaired phase separation and have been implicated in neurodegenerative and rare genetic disorders (10–12).

CREBBP (CBP) and its paralog EP300 (p300) are transcriptional coactivators with histone acetyltransferase (HAT) activity that contain extensive IDRs. In the p300/CBP coactivator family, enzymatic activity is regulated by the acetylation of the autoinhibitory loop. Under basal conditions, an α-helix located within the N-terminal portion of the ID4 region is positioned near the catalytic site and is thought to contribute to autoinhibition. Upon interacting with transcription factors or conformational rearrangement, the subsequent dissociation of this helix from the catalytic site activates HAT (13–15).

Moreover, EP300 forms cocondensates with transcription factors through IDR-mediated interactions and enables the recruitment of transcriptional regulators, such as BRD4 to regulate gene expression (13,14,16). Similar mechanisms supposedly operate in CREBBP. However, the detailed molecular basis remains unclear.

Reduced CREBBP HAT activity or haploinsufficiency causes Rubinstein–Taybi syndrome (RSTS) (17). In contrast, missense and in-frame indel variants located within the C-terminal ZZ-type zinc finger (ZZ), transcriptional adapter zinc-binding 2 (TAZ2), and fourth intrinsically disordered linker (CBP-ID4) domains of *CREBBP* are responsible for a distinct clinical entity known as Menke– Hennekam syndrome (MKHK) (18,19). In particular, patients harboring variants in the ID4 domain exhibit a characteristic DNA methylation episignature, although the molecular mechanism underlying this phenotype remains unknown (19,20). These observations suggest that distinct variants within the same gene may cause the disease through fundamentally different molecular mechanisms. However, the molecular basis by which ID4-domain variants in *CREBBP* cause the unique clinical and epigenetic features of MKHK remains largely unknown. Elucidating the variant-specific molecular effects of CREBBP is therefore critical for a more comprehensive understanding of the pathogenesis of MKHK.

In the current study, we identified the recurrent *CREBBP* missense variant p.(Arg1868Trp) and novel splice variant r.5963_5969del,p.(Cys1723Thrfs*19) in two patients clinically diagnosed with MKHK. Thereafter, we investigated how these variants affected HAT activity and nuclear phase behavior to obtain a deep understanding of the molecular pathogenesis of MKHK.

## Results

### Clinical findings

The proband of family 1 (F1) was a girl with severe developmental delay and substantial growth impairment. She presented with bilateral renal hypoplasia, sensorineural hearing loss, contractures of the elbows, knees, and hip joints, bilateral fifth-finger clinodactyly and syndactyly of the right second and third fingers. Her facial characteristics included frontal bossing, hypertelorism, deep-set eyes, short palpebral fissures, sparse eyebrows, a prominent nasal root, hypoplastic alae nasi, a flat nasal bridge, thin lips, abnormal ear morphology, and sparse scalp hair (Table 1, Fig. S1A). The proband of family 2 (F2) was a girl who had been diagnosed with congenital heart defects, including ventricular septal defect and aortic stenosis, cerebellar hypoplasia, and fetal growth restriction during the fetal period. Postnatally, she exhibited developmental delays and severe growth impairment, as well as a horseshoe kidney and syndactyly of the left third to fifth fingers. Facial features included frontal bossing, upslanted palpebral fissures, ptosis/narrow palpebral fissures, hypoplasia of the nasal root, a short columella, anteverted nares, and a long philtrum (Table 1 and Fig. S1B). Representative clinical and molecular features of the two individuals included in this study and previously reported as patients with MKHK having CREBBP exon 31 variants are summarized in Table 1.

**Table 1.**
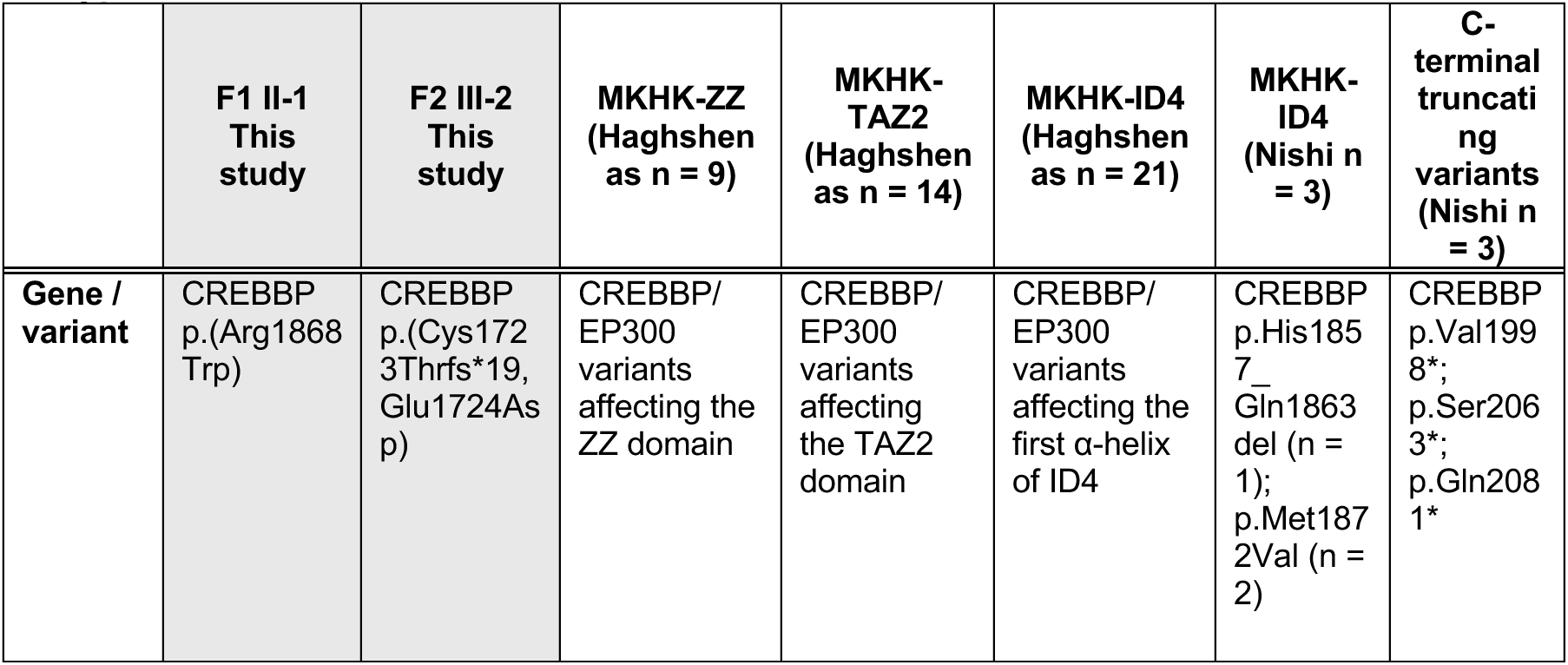
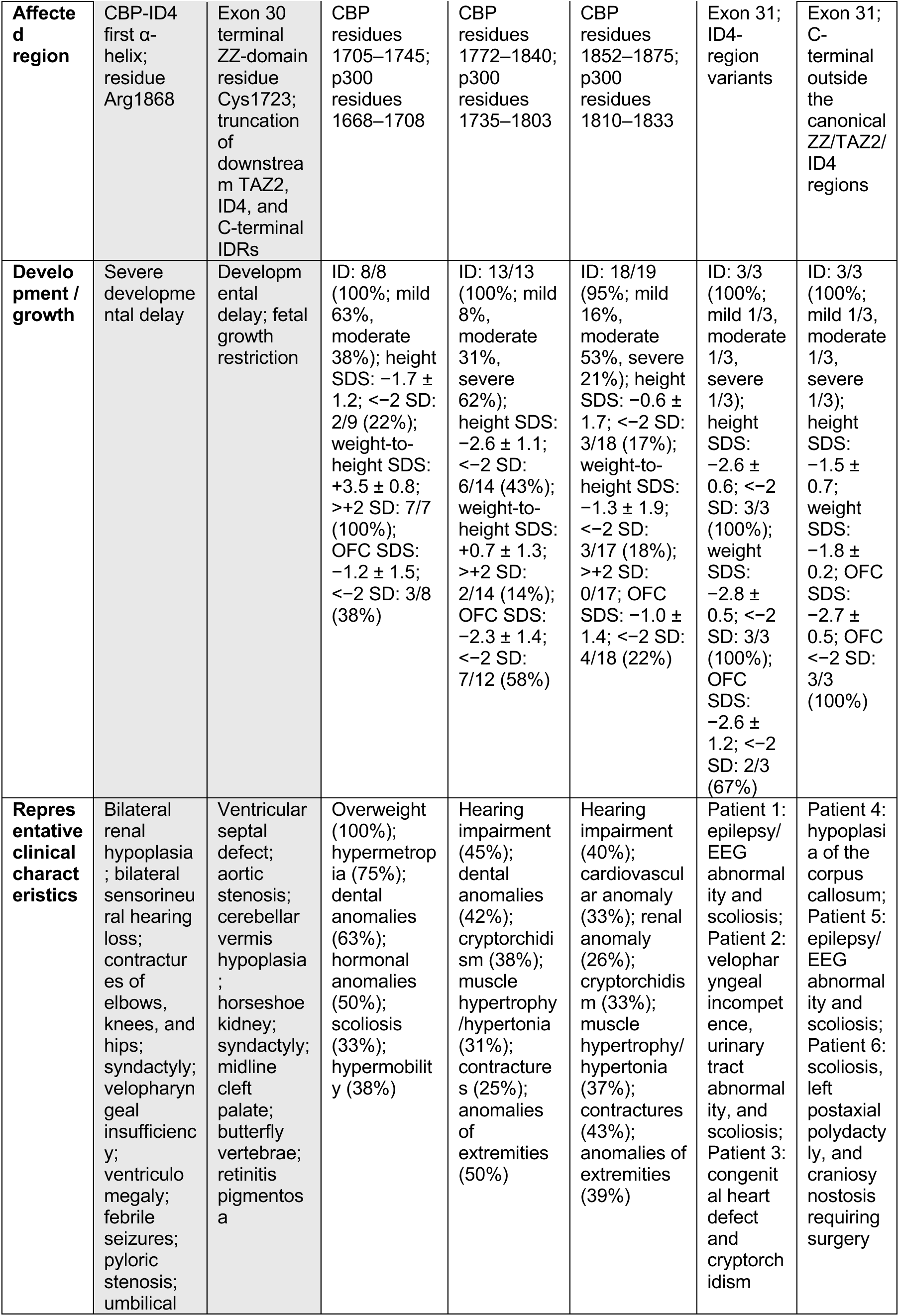
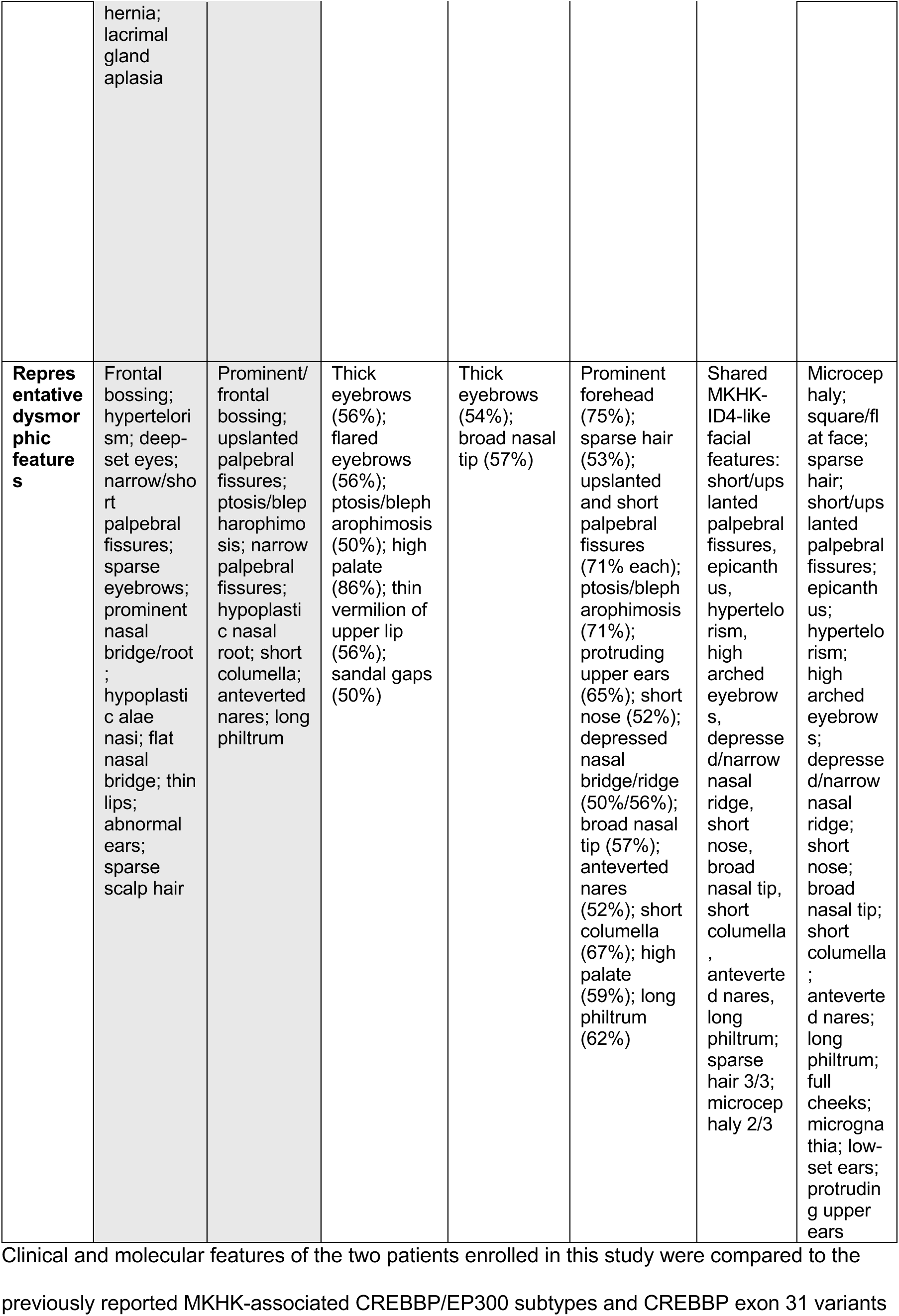
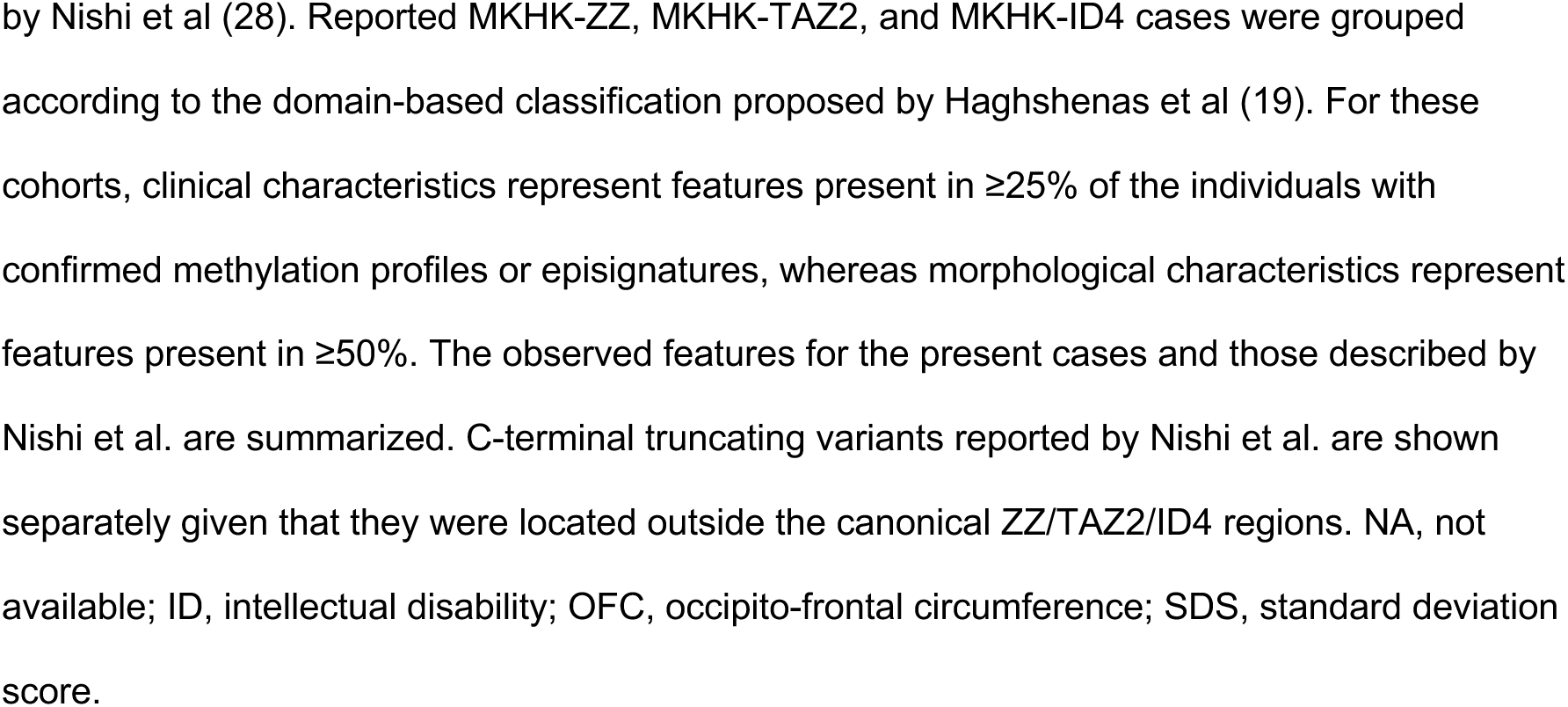
Clinical and molecular comparison of the current cases with reported MKHK subtypes.

|  | F1 II-1<br>This study | F2 III-2<br>This study | MKHK-ZZ<br>(Haghshen<br>as n = 9) | MKHK-TAZ2<br>(Haghshen<br>as n = 14) | MKHK-ID4<br>(Haghshen<br>as n = 21) | MKHK-ID4<br>(Nishi n = 3) | C-terminal truncating variants<br>(Nishi n = 3) |
| --- | --- | --- | --- | --- | --- | --- | --- |
| **Gene / variant** | CREBBP p.(Arg1868 Trp) | CREBBP p.(Cys172 3Thrfs*19, Glu1724Asp) | CREBBP/EP300 variants affecting the ZZ domain | CREBBP/EP300 variants affecting the TAZ2 domain | CREBBP/EP300 variants affecting the first α-helix of ID4 | CREBBP p.His185 7_ Gln1863 del (n = 1); p.Met187 2Val (n = 2) | CREBBP p.Val199 8*; p.Ser206 3*; p.Gln208 1* |
| **Affected region** | CBP-ID4 first α-helix; residue Arg1868 | Exon 30 terminal ZZ-domain residue Cys1723; truncation of downstream TAZ2, ID4, and C-terminal IDRs | CBP residues 1705–1745; p300 residues 1668–1708 | CBP residues 1772–1840; p300 residues 1735–1803 | CBP residues 1852–1875; p300 residues 1810–1833 | Exon 31; ID4-region variants | Exon 31; C-terminal outside the canonical ZZ/TAZ2/ID4 regions |
| **Development / growth** | Severe developmental delay | Developmental delay; fetal growth restriction | ID: 8/8 (100%; mild 63%, moderate 38%); height SDS: $-1.7 \pm 1.2$; $<-2$ SD: 2/9 (22%); weight-to-height SDS: $+3.5 \pm 0.8$; $>+2$ SD: 7/7 (100%); OFC SDS: $-1.2 \pm 1.5$; $<-2$ SD: 3/8 (38%) | ID: 13/13 (100%; mild 8%, moderate 31%, severe 62%); height SDS: $-2.6 \pm 1.1$; $<-2$ SD: 6/14 (43%); weight-to-height SDS: $+0.7 \pm 1.3$; $>+2$ SD: 2/14 (14%); OFC SDS: $-2.3 \pm 1.4$; $<-2$ SD: 7/12 (58%) | ID: 18/19 (95%; mild 16%, moderate 53%, severe 21%); height SDS: $-0.6 \pm 1.7$; $<-2$ SD: 3/18 (17%); weight-to-height SDS: $-1.3 \pm 1.9$; $<-2$ SD: 3/17 (18%); $>+2$ SD: 0/17; OFC SDS: $-1.0 \pm 1.4$; $<-2$ SD: 4/18 (22%) | ID: 3/3 (100%; mild 1/3, moderate 1/3, severe 1/3); height SDS: $-2.6 \pm 0.6$; $<-2$ SD: 3/3 (100%); weight SDS: $-2.8 \pm 0.5$; $<-2$ SD: 3/3 (100%); OFC SDS: $-2.6 \pm 1.2$; $<-2$ SD: 2/3 (67%) | ID: 3/3 (100%; mild 1/3, moderate 1/3, severe 1/3); height SDS: $-1.5 \pm 0.7$; weight SDS: $-1.8 \pm 0.2$; OFC SDS: $-2.7 \pm 0.5$; OFC $<-2$ SD: 3/3 (100%) |
| **Representative clinical characteristics** | Bilateral renal hypoplasia; bilateral sensorineural hearing loss; contractures of elbows, knees, and hips; syndactyly; velopharyngeal insufficiency; ventriculomegaly; febrile seizures; pyloric stenosis; umbilical hernia;<br>lacrimal<br>gland<br>aplasia | Ventricular septal defect; aortic stenosis; cerebellar vermis hypoplasia; horseshoe kidney; syndactyly; midline cleft palate; butterfly vertebrae; retinitis pigmentosa | Overweight (100%); hypermetropia (75%); dental anomalies (63%); hormonal anomalies (50%); scoliosis (33%); hypermobility (38%) | Hearing impairment (45%); dental anomalies (42%); cryptorchidism (38%); muscle hypertrophy/hypertonia (31%); contractures (25%); anomalies of extremities (50%) | Hearing impairment (40%); cardiovascular anomaly (33%); renal anomaly (26%); cryptorchidism (33%); muscle hypertrophy/hypertonia (37%); contractures (43%); anomalies of extremities (39%) | Patient 1: epilepsy/EEG abnormality and scoliosis; Patient 2: velopharyngeal incompetence, urinary tract abnormality, and scoliosis; Patient 3: congenital heart defect and cryptorchidism | Patient 4: hypoplasia of the corpus callosum; Patient 5: epilepsy/EEG abnormality and scoliosis; Patient 6: scoliosis, left postaxial polydactyly, and craniosynostosis requiring surgery |
| **Representative dysmorphic features** | Frontal bossing; hypertelorism; deep-set eyes; narrow/short palpebral fissures; sparse eyebrows; prominent nasal bridge/root; hypoplastic alae nasi; flat nasal bridge; thin lips; abnormal ears; sparse scalp hair | Prominent/frontal bossing; upslanted palpebral fissures; ptosis/blepharophimosis; narrow palpebral fissures; hypoplastic nasal root; short columella; anteverted nares; long philtrum | Thick eyebrows (56%); flared eyebrows (56%); ptosis/blepharophimosis (50%); high palate (86%); thin vermilion of upper lip (56%); sandal gaps (50%) | Thick eyebrows (54%); broad nasal tip (57%) | Prominent forehead (75%); sparse hair (53%); upslanted and short palpebral fissures (71% each); ptosis/blepharophimosis (71%); protruding upper ears (65%); short nose (52%); depressed nasal bridge/ridge (50%/56%); broad nasal tip (57%); anteverted nares (52%); short columella (67%); high palate (59%); long philtrum (62%) | Shared MKHK-ID4-like facial features: short/upslanted palpebral fissures, epicanthus, hypertelorism, high arched eyebrows, depressed/narrow nasal ridge, short nose, broad nasal tip, short columella, anteverted nares, long philtrum; sparse hair 3/3; microcephaly 2/3 | Microcephaly; square/flat face; sparse hair; short/upslanted palpebral fissures; epicanthus; hypertelorism; high arched eyebrows; depressed/narrow nasal ridge; short nose; broad nasal tip; short columella; anteverted nares; long philtrum; full cheeks; micrognathia; low-set ears; protruding upper ears |

Clinical and molecular features of the two patients enrolled in this study were compared to the previously reported MKHK-associated CREBBP/EP300 subtypes and CREBBP exon 31 variants
by Nishi et al (28). Reported MKHK-ZZ, MKHK-TAZ2, and MKHK-ID4 cases were grouped according to the domain-based classification proposed by Haghshenas et al (19). For these cohorts, clinical characteristics represent features present in $\geq 25\%$ of the individuals with confirmed methylation profiles or episignatures, whereas morphological characteristics represent features present in $\geq 50\%$ . The observed features for the present cases and those described by Nishi et al. are summarized. C-terminal truncating variants reported by Nishi et al. are shown separately given that they were located outside the canonical ZZ/TAZ2/ID4 regions. NA, not available; ID, intellectual disability; OFC, occipito-frontal circumference; SDS, standard deviation score.

### Identification of recurrent and novel *CREBBP* variants in two patients with MKHK

In family F1, we identified a recurrent de novo missense variant in MKHK (*CREBBP*: NM_004380.3: c.5602C>T,p.(Arg1868Trp) [R1868W]) (Fig. S1A) (18–20). In family F2, we identified a novel de novo splice variant located at the end of exon 30 (NC_000016.10:g.3731192C>G) (Fig. S1B).

### Splicing analysis of the variant located at the end of exon 30

To determine the effects of the exon 30 terminal variant on splicing, RT-PCR was performed using peripheral blood obtained from the proband (III-2). TA cloning analysis of seven independent clones revealed two aberrant transcript species. Among the seven clones, three of the contained a 7-bp deletion, resulting in a frameshift transcript (r.5963_5969del), whereas one retained normal splicing but carried the nucleotide substitution transcript (r.5969g>c) (*CREBBP*: NM_004380.3: r.[5963_5969del,5969g>c], p.(Cys1723Thrfs*19, Glu1724Asp) [C1723fs, E1724D]) (Fig. S2A). The remaining clones corresponded to wild-type transcripts. Hereinafter, p.(Cys1723Thrfs*19) is referred to as C1723fs.

### Expression of the CREBBP protein from the C1723 frameshift variant

Thereafter, we evaluated CREBBP protein expression in patient-derived lymphoblastoid cell lines (LCLs). SDS-PAGE and Western blot analysis revealed that aside from wild-type CREBBP (∼300 kDa), a truncated CREBBP protein of approximately 190 kDa was identified in the F2 proband (III-2) (Fig. S2B). The C1723fs transcript escapes nonsense-mediated mRNA decay (NMD) and is expressed as a truncated protein product.

### HAT activity of C1723fs and R1868W CREBBP protein

The ID4 region of CREBBP (CBP-ID4) is an IDR spanning amino acids 1851–2066, portions of which (residues 1852–1875 and 1951–1978) have been reported to adopt partial α-helical conformations (Figure 1A) (21). In EP300, the N-terminal α-helix within ID4 is positioned near the catalytic site of the HAT domain, and the dissociation of this helix from the catalytic site upon interaction with transcription factors or BRD4-NUT promotes enzymatic activation (15).

**Figure 1.**
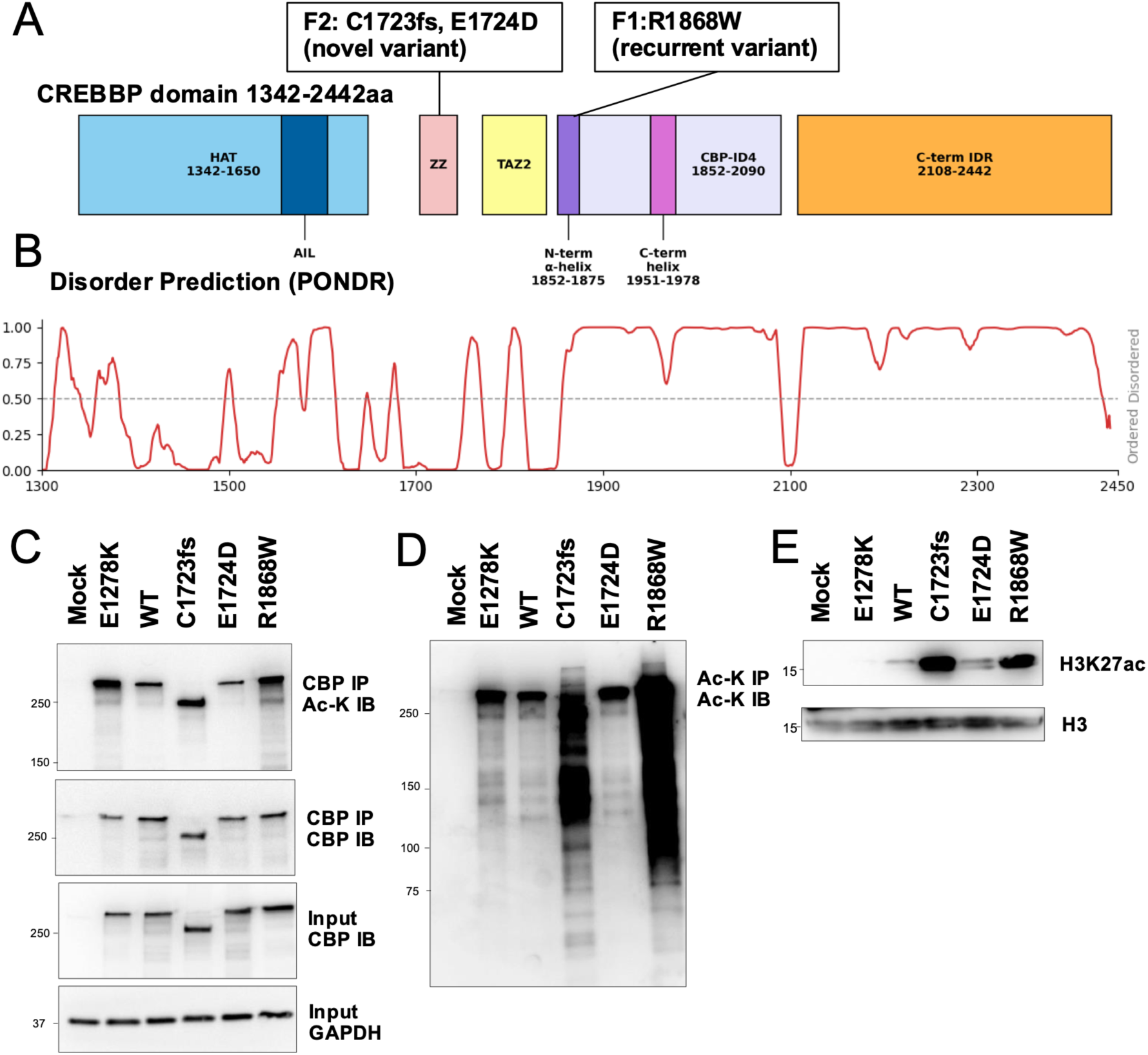
*CREBBP* variants in Menke–Hennekam syndrome and their biochemical characterization. **(A)** Domain architecture of CREBBP and locations of pathogenic variants identifed in individuals clinically diagnosed with MKHK (F1 II-1 and F2 III-2). **(B)** Prediction of intrinsically disordered regions (IDRs) within CREBBP using PONDR (22). **(C)** Analysis of CREBBP autoacetylation. CREBBP was immunoprecipitated using an anti-CBP antibody, followed by immunoblotting with anti–acetyl-lysine (CBP IP, Ac-K IB) or anti-CBP (CBP IP, CBP IB) antibodies. **(D)** Analysis of nonhistone protein acetylation. Acetylated proteins were immunoprecipitated using acetyl-lysine-binding beads and detected by immunoblotting with anti-acetyl-lysine antibody (Ac-K IP, Ac-K IB). **(E)** Analysis of histone acetylation. Histone extracts were subjected to immunoblotting using antibodies against H3 and H3K27ac.

A previously reported RSTS causative variant (E1278K), which has been shown to impair HAT activity (23) in assays using an isolated CBP HAT-domain fragment, was included for comparison. Given that the variants detected in the current study were predicted to alter the regulation of CREBBP HAT activity, we investigated the functional outcomes. Accordingly, HaloTag-fused CREBBP constructs (wild-type, R1868W, C1723fs, E1724D, and E1278K) were transiently expressed in HEK293 cells, followed by CREBBP immunoprecipitation, acetyl-lysine pull-down assays, and histone extraction analyses.

Both R1868W and C1723fs exhibited increased acetyl-lysine signals in CREBBP immunoprecipitates, enhanced acetylation of nonhistone proteins, and elevated H3K27ac levels compared to wild-type CREBBP (Figure 1C–E). In contrast, E1278K showed increased acetyl-lysine signals in CREBBP immunoprecipitates but did not exhibit increased nonhistone protein acetylation or elevated H3K27ac levels (Figure 1C–E) (23).

### MKHK causative variants disrupt liquid–liquid phase separation in divergent ways

Previous studies have shown that deletion of the region corresponding to the N-terminal α-helix within the ID4 domain of EP300, a homolog of CREBBP, induces nuclear condensate formation, accompanied by higher HAT activity (15). Moreover, evidence suggests that the C-terminal region of CREBBP constitutes an IDR (Figure 1B) containing glutamine-rich sequences and a polyQ (23). Loss of the C-terminal glutamine-rich IDR has been associated with impaired phase separation (23).

To investigate the effects of MKHK with *CREBBP* pathogenic variants (R1868W and C1723fs) on nuclear condensate formation, HaloTag-fused full-length human CREBBP constructs (wild-type, R1868W, and C1723fs) were transiently expressed in COS7 cells and visualized using a confocal laser-scanning microscope. Wild-type CREBBP formed punctate nuclear condensates, R1868W formed enlarged condensates, and C1723fs exhibited diffuse nuclear localization (Figure 2A).

**Figure 2.**
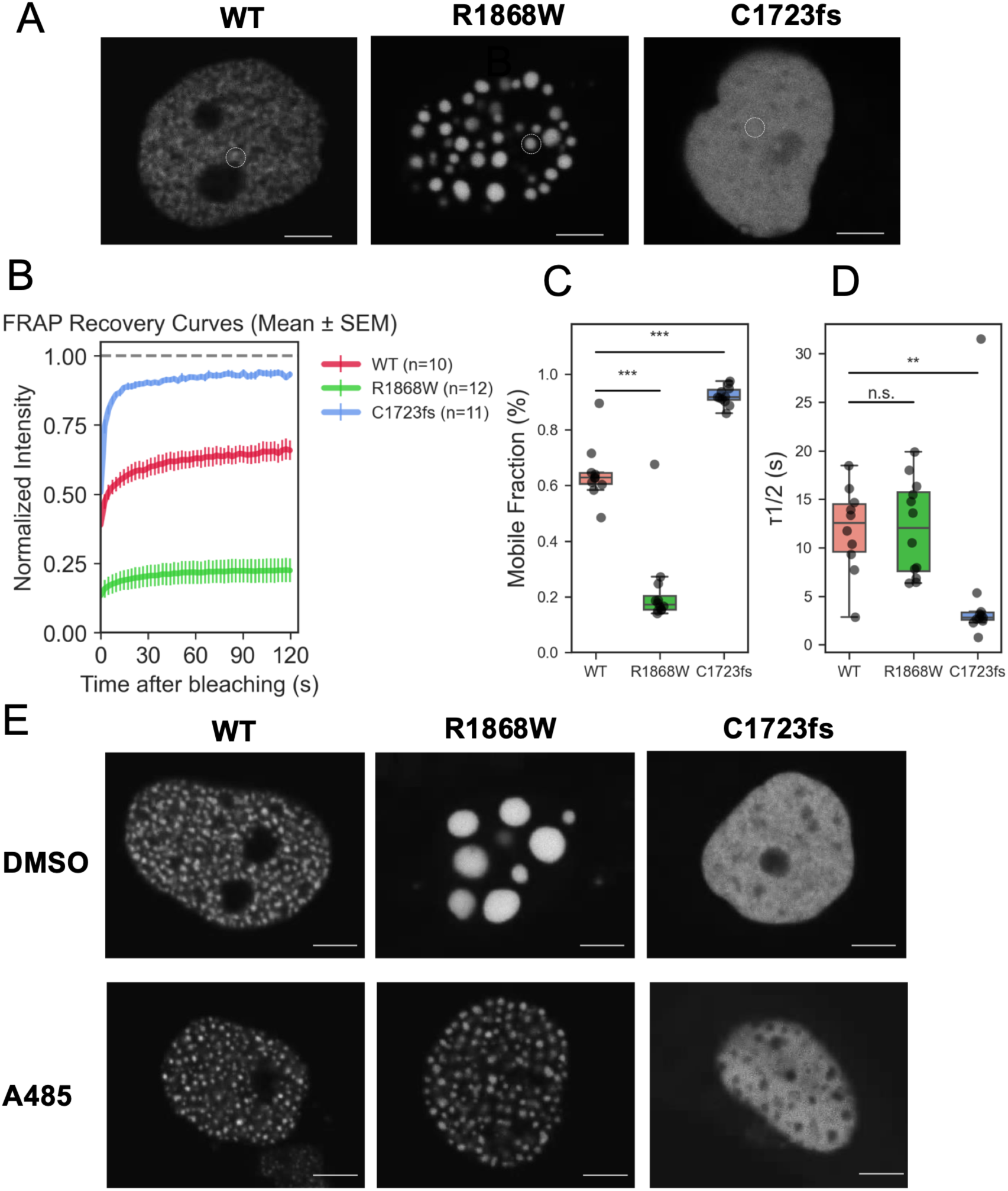
Altered condensate dynamics of *CREB-binding protein* (*CREBBP)* variants revealed through fluorescence recovery after photobleaching (FRAP) analysis. **(A)** Representative images of COS7 cells transiently expressing HaloTag-fused CREBBP (WT, R1868W, and C1723fs) following labeling with TMR ligand and analyzed via FRAP using a FV3000 confocal microscope. Images obtained prior to photobleaching are shown. Dashed circles indicate bleached regions. Scale bar, 5 µm. **(B)** Normalized fluorescence recovery curves for each construct before and after photobleaching. Fluorescence intensities were normalized to pre-bleach levels, with photobleaching defined as *t* = 0. Solid lines represent mean normalized fluorescence intensity at each time point, and error bars indicate the standard error of the mean (SEM). n = 10, 12, and 11 bleached regions from independent cells for WT, R1868W, and C1723fs, respectively. **(C)** Quantification of the mobile fraction for each construct. **(D)** Quantification of fluorescence recovery half-time (τ) for each construct. **(E)** Representative nuclear images of COS7 cells expressing HaloTag-fused CREBBP (WT, R1868W, and C1723fs) after treatment with DMSO or A485 for 1 hour, following TMR ligand labeling. Scale bar, 5 µm.

To evaluate condensate dynamics, fluorescence recovery after photobleaching (FRAP) analysis was performed. Wild-type CREBBP exhibited punctate nuclear condensates with a mobile fraction of 0.64 ± 0.11 (mean ± SD) fluorescence recovery and half-recovery times (τ1/2) of ∼12 s, consistent with a dynamic molecular exchange. In contrast, R1868W showed markedly reduced fluorescence recovery (mobile fraction, R1868W vs. WT: 0.22 ± 0.15; P = 0.000684), indicating a severely restricted molecular exchange between the condensates and surrounding nucleoplasm. Conversely, C1723fs displayed diffuse nuclear localization and revealed high fluorescence recovery (C1723fs vs. WT: 0.92 ± 0.03; P = 0.000575), together with significantly shortened τ1/2 values than WT (P = 0.0165), consistent with behavior approaching free nuclear diffusion (Figure 2B–D).

Thereafter, to investigate the role of HAT activity in nuclear condensate formation, cells expressing CREBBP were treated with the selective p300/CBP HAT inhibitor A485. Accordingly, wild-type CREBBP formed punctate condensates under both DMSO and A485 treatment conditions. In contrast, R1868W formed large condensates measuring 3.08 ± 0.97 µm in diameter (mean ± SD; n = 8 condensates) under DMSO treatment, which reduced in size after A485 treatment. C1723fs exhibited diffuse nuclear localization under both conditions (Figure 2E).

### Aberrant condensates formed by R1868W exhibit restored dynamics upon HAT inhibition

We subsequently compared the FRAP dynamics of R1868W condensates under DMSO and A485 conditions. Notably, A485 treatment significantly improved fluorescence recovery (mobile fraction, A485 vs. DMSO: 0.522 ± 0.068 vs 0.347 ± 0.088 (mean ± SD); P = 0.000224) and reduced the apparent condensate size (Figure 3A–C).

**Figure 3.**
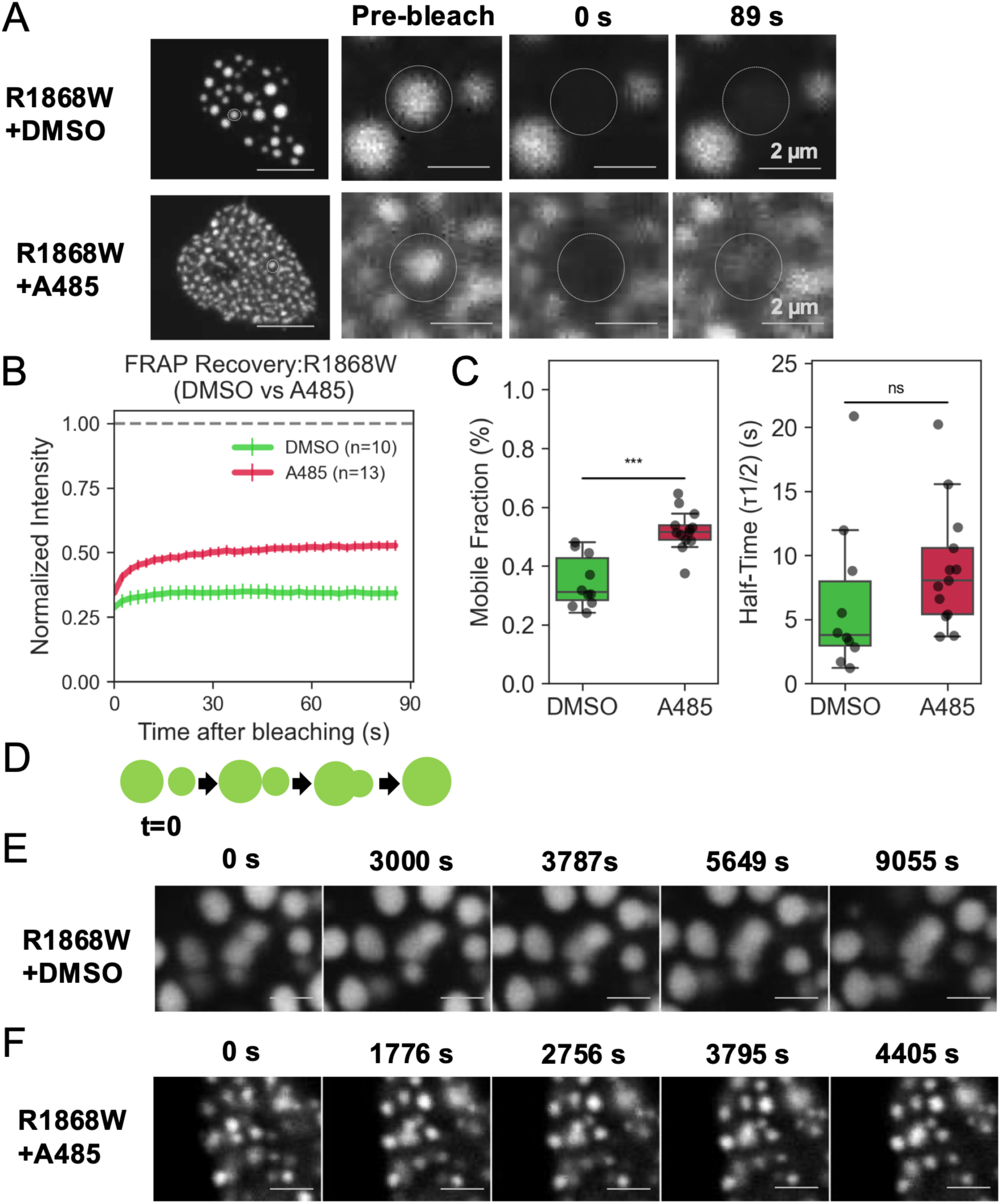
A485 restores the dynamic properties of CREB-binding protein (CREBBP) R1868W condensates. **(A)** FRAP analysis of COS7 cells transiently expressing HaloTag-fused CREBBP R1868W, following TMR ligand labeling and treatment with DMSO or A485 for 1 hour. Representative pre-bleach images are shown on the left, with dashed circles indicating the bleached regions (scale bar, 5 µm). Enlarged views of the bleached regions at pre-bleach, *t* = 0, and *t* = 89 s are displayed (scale bar, 2 µm). **(B)** Normalized fluorescence recovery curves for R1868W condensates, treated with DMSO or A485. Fluorescence intensities were normalized to pre-bleach levels, with photobleaching defined as *t* = 0. Solid lines represent the mean normalized fluorescence intensity at each time point, whereas error bars indicate the standard error of the mean (SEM). For FRAP analysis, n = 10 and 13 condensates for DMSO and A485, respectively. **(C)** Quantification of the mobile fraction and fluorescence recovery half-time (τ) under each condition. **(D)** Schematic illustration of the time-lapse analysis of condensate fusion and relaxation in R1868W-expressing cells, treated with DMSO or A485. **(E)** , **(F)** Representative time-lapse images showing fusion and relaxation dynamics of nuclear condensates. Enlarged views of individual condensates undergoing fusion and relaxation at indicated time points are displayed (scale bar, 1 µm).

To further evaluate condensate behavior, live-cell time-lapse imaging was performed. Under DMSO conditions, dumbbell-shaped structures associated with condensate fusion intermediates were observed; however, clear relaxation into spherical structures was not detected within the observation period (150 min). In contrast, under A485 treatment conditions, smaller and more numerous condensates were detected, and multiple fusion and relaxation events were recorded during imaging (Figure 3D–F).

### Aberrant condensates formed by R1868W recruit BRD4 in an acetylation-dependent manner

Co-condensation between transcription factors and EP300 activates HAT and recruits BRD4 to chromatin through BRD4 bromodomain-dependent recognition of acetylated lysine residues (14,16). We therefore investigated whether aberrant condensates formed by CREBBP R1868W similarly recruit BRD4 in an acetylation-dependent manner.

Immunostaining revealed the accumulation of H3K27ac around R1868W condensates (Fig. S4). To quantify BRD4 enrichment around condensates, radial fluorescence intensity distribution analysis was conducted by normalizing distances from the condensate center to its radius (Figure 4B). Accordingly, JQ1, a BET bromodomain inhibitor, reduced BRD4 fluorescence intensity surrounding R1868W condensates compared to DMSO-treated cells, indicating that BRD4 recruitment to aberrant condensates depends on bromodomain-mediated recognition of acetylated chromatin (Figure 4A, 4C).

**Figure 4.**
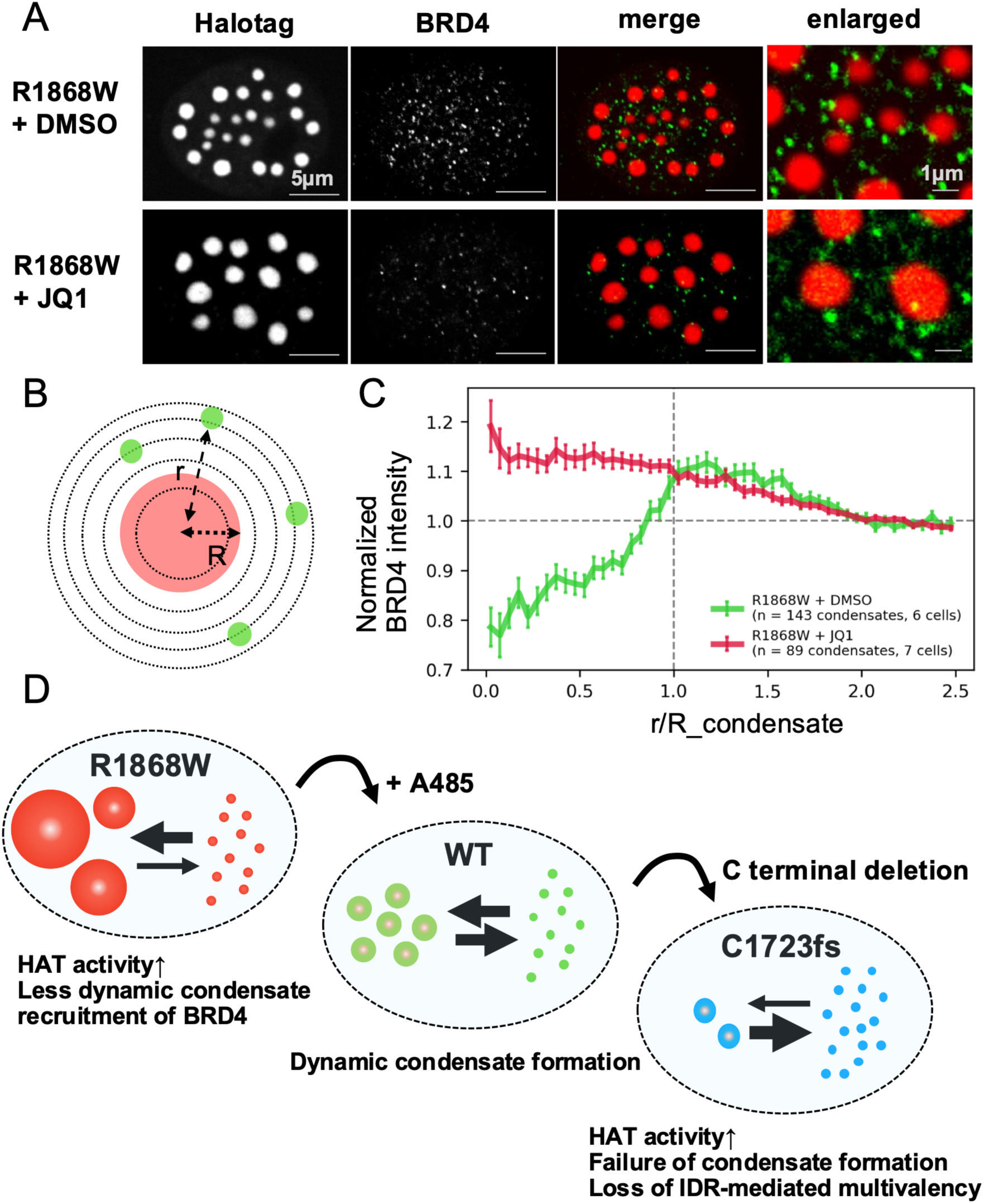
BRD4 enrichment around CREB-binding protein (CREBBP) condensates depending on acetylation and being altered by BET inhibition. **(A)** Representative single optical sections of R1868W-expressing cells treated with DMSO or JQ1. COS7 cells transiently expressing HaloTag-fused CREBBP R1868W were labeled with TMR ligand, treated with DMSO or the BET inhibitor JQ1 for 1 hour, and subjected to immunostaining with an anti-BRD4 antibody. From left to right: HaloTag, BRD4, and merged images (scale bar, 5 µm). The rightmost panels show enlarged merged images of condensates (scale bar, 1 µm). **(B)** Schematic of the radial distribution analysis. To assess acetylation-dependent recruitment of BRD4 to nuclear condensates, BRD4 fluorescence intensity was quantified as a function of distance from the condensate center, normalized to the condensate radius (r/R). **(C)** Radial distribution profiles of BRD4 intensity as a function of normalized distance (r/R_condensate). Fluorescence intensities were normalized to the mean nuclear intensity. Solid lines represent mean normalized intensity at each distance, and error bars indicate the standard error of the mean (SEM). A total of 143 condensates from 6 DMSO-treated cells and 89 condensates from 7 JQ1-treated cells were analyzed. **(D)** Model summarizing the relationship between CREBBP variants and LLPS behavior. Wild-type CREBBP forms dynamic punctate nuclear condensates. The R1868W variant forms aberrant condensates with reduced dynamics and increased HAT activity, accompanied by enhanced BRD4 enrichment. In contrast, the C1723fs variant lacks the C-terminal IDR, thereby impairing LLPS and resulting in a diffuse nuclear distribution. Inhibition of HAT activity by A485 partially restored the dynamic properties of R1868W condensates.

## Discussion

The current study demonstrated that pathogenic *CREBBP* variants responsible for MKHK are both characterized by pronounced HAT activity while disrupting LLPS through distinct mechanisms.

The recurrent MKHK-ID4 variant R1868W formed aberrant condensates with reduced fluidity, at least partly in association with increased HAT activity. FRAP and live-cell time-lapse analyses showed decreased molecular exchange and delayed relaxation into spherical structures following fusion, consistent with a less dynamic condensate state. Furthermore, these condensates modulated the transcriptional regulator BRD4 in an acetylated histone–dependent manner, suggesting that the spatial organization and dynamics of transcriptional regulatory compartments can be altered. Simultaneously, R1868W can also disrupt the N-terminal α-helical structure within the CBP-ID4 region, thereby altering the flexibility and multivalent interactions of the IDR. Thus, a single mechanism alone may not explain the altered phase behavior induced by R1868W. In contrast, although the C1723fs variant also exhibited elevated HAT activity, the deletion of the C-terminal IDR promoted the loss of glutamine-rich sequences implicated in LLPS, likely interfering with the phase separation itself. Consequently, CREBBP failed to form nuclear condensates, potentially destabilizing the scaffold, required for transcriptional complex assembly, and thereby modulating the transcriptional regulatory environment through a mechanism, distinct from that of R1868W.

In the present study, we did not perform transcriptome-wide analyses, such as RNA-seq or chromatin profiling, and therefore the precise relationship between altered condensate properties and downstream transcriptional programs or phenotypic manifestations remains to be investigated. Nevertheless, the abnormal BRD4 expression and altered condensate dynamics observed in the present study support the possibility that disruption of transcriptional regulatory environments contributes to disease pathogenesis of *CREBBP* variants in MKHK.

Notably, although these two variants shared increased HAT activity, they produced contrasting LLPS phenotypes, namely “reduced condensate fluidity” and “failure of phase separation.” These findings suggest that a shared enzymatic abnormality can differentially alter condensate material properties and thereby exert distinct effects on transcriptional regulatory environments. Thus, the molecular pathogenesis of MKHK can be understood not solely as an enzymatic disorder but also as a disorder of condensate material properties (Figure 4D).

Importantly, treatment with the HAT inhibitor A485 partially restored condensate fluidity and reduced condensate size in R1868W-expressing cells. These findings suggest that aberrant condensate properties are, at least in part, pharmacologically reversible, raising the possibility that modulating HAT activity can serve as a potential therapeutic strategy. In contrast, for variants such as C1723fs that impair condensate formation itself, simple enzymatic inhibition alone may be insufficient, highlighting the need for disease subtype-specific therapeutic approaches. The C1723fs variant identified in family F2 exhibited characteristic MKHK facial features while lacking the characteristic RSTS facial or limb features. Unlike previously reported MKHK variants, which are primarily missense variants or small in-frame deletions within the ZZ, TAZ2, and ID4 domains, C1723fs is characterized by extensive loss of the C-terminal IDR. These findings suggest that the clinical manifestations of MKHK may emerge not only from localized dysfunction of specific domains but also from broader alterations affecting phase behavior and IDR-dependent interactions of CREBBP.

DNA methylation analysis involving the previously reported MKHK-ID4-specific methylation probes demonstrated that the two MKHK cases (R1868W and C1723fs) clustered together and were distinguished from RSTS cases and healthy controls (25) (Fig. S3). These findings suggest that the C1723fs-associated methylation profile shares epigenomic features with *CREBBP* variants in MKHK.

However, the MKHK-associated methylation probes employed in our research were originally developed using previously reported ID4-related variants, and the corresponding training datasets and raw methylation data had not been publicly released (19,20). Furthermore, the number of pathogenic reference samples available for comparative analysis remains limited. Therefore, it is still unclear whether the methylation profile observed in the C1723fs case represents the same characteristic MKHK-ID4 episignature as previously reported cases or a distinct epigenomic pattern. Additional studies including a larger cohort of patients with MKHK are needed to determine whether the methylation profile associated with C1723fs represents the canonical MKHK-ID4 episignature or a distinct epigenomic pattern (19,20).

More broadly, our findings are consistent with emerging concepts suggesting that disease-causing variants affecting enzymatic activity or IDR composition can perturb the formation and material properties of LLPS-mediated molecular assemblies, thereby contributing to disease pathogenesis (10–12). Within this framework, alterations in condensate formation, dynamics, and material properties can disrupt normal molecular functions and contribute to the development of inherited diseases.

Taken together, our study provides a conceptual framework in which the molecular pathogenesis of MKHK would be regarded as a disorder whose mechanisms involve both enzymatic dysregulation and altered condensate material properties.

## Materials and Methods

### Ethical statement

This study was approved by the Ethics Committee of the National Center for Child Health and Development (Approval No. 2020-326). Written informed consent was obtained from the parents or legal guardians of the probands and from participating family members for the clinical evaluation, genetic analysis, use of established patient-derived cell lines and publication of anonymized clinical information and genetic information. Informed consent for the publication of clinical photographs was also obtained from the parents or legal guardians of each proband.

### Whole-exome sequencing and filtering analysis

Whole-exome sequencing (WES) and filtering analysis were conducted in the patients from in Families 1 and 2 and their parents, with minor modifications to the previously described procedure (26).

In brief, genomic DNA was extracted from peripheral blood leukocytes. For family F1, exome-enriched libraries were prepared using the SureSelect Human All Exon V6 kit (Agilent Technologies, Santa Clara, CA, USA) and sequenced on a HiSeq 2500 platform (Illumina, San Diego, CA, USA). For family F2, libraries were prepared using the Twist Comprehensive Exome with Mitochondrial DNA Panel Spike-In (Twist Bioscience, San Francisco, CA, USA) and sequenced on a NovaSeq 6000 platform (Illumina, San Diego, CA, USA). Sequencing reads were aligned to the GRCh38 reference genome with using the Burrows–Wheeler Aligner. Variant calling and annotation were conducted with the CompStor Genomic Analysis Platform (OmniTier). Variants were filtered according to inheritance pattern and allele frequency, after which variants with allele frequencies <0.01 in gnomAD v4.1.0, ToMMo 54KJPN, and our in-house WES database were extracted for further analysis.

### Transcript sequencing

Total RNA was extracted from peripheral blood obtained from the F2 proband (III-2), and reverse transcription was performed using the PrimeScript 1st Strand cDNA Synthesis Kit (Takara Bio, Shiga, Japan).

To evaluate the splice variant identified via WES, primers spanning CREBBP exons 29–31 were designed (Table S2), after which PCR amplification was performed. PCR products were then subjected to TA cloning, followed by Sanger sequencing of individual clones (n = 7). The proportion of each transcript was estimated based on clone counts.

### Establishment and culture of lymphoblastoid cell lines

LCLs obtained from F2 (III-2) were established by SRL Inc. (Tokyo, Japan). Peripheral blood mononuclear cells were isolated and immortalized using the culture supernatant from the EBV-producing marmoset B95-8 cell line.

Cells were cultured in RPMI-1640 medium supplemented with 10% fetal bovine serum and 100 U/mL penicillin–streptomycin (Gibco, Thermo Fisher Scientific, Waltham, MA, United States).

### Western blotting

Control and F2 III-2–derived LCLs were lysed in RIPA buffer (#16488, Nacalai Tesque, Kyoto, Japan) with cOmplete™ Protease Inhibitor Cocktail (Roche Diagnostics, Basel, Switzerland) for 30 min on ice, centrifuged at 12,000 rpm at 4 °C, further mixed with SDS sample buffer, separated on 4%–15% Mini-PROTEAN® TGX™ gels (Bio-Rad, Hercules, CA, United States) at 120 V for 60 min, and then transfered to PVDF membranes at 100 V for 60 min.

Membranes were incubated overnight at 4°C with either CBP (D6C5) Rabbit Monoclonal Antibody (1:2,000; #7389, Cell Signaling Technology, Danvers, MA, United States) or Anti-GAPDH (14C10) Rabbit mAb (1:2,000; #2118, Cell Signaling Technology). HRP-conjugated anti-rabbit IgG secondary antibody (1:2,000; #7074, Cell Signaling Technology) was applied for 90 min at room temperature.

### Episignature analysis

Genomic DNA extracted from peripheral blood leukocytes of the two probands (F1 II-1 and F2 III-2) and two in-house healthy controls (one adult female and one female child) was subjected to bisulfite conversion and analyzed using the Illumina Infinium Methylation EPIC BeadChip (EPIC) array (Illumina, San Diego, CA, United States).

Publicly available methylation datasets from 55 healthy children (23 males and 32 females, aged 2–15 years) included in GSE180534 and GSE193730 were analyzed as controls. Moreover, DNA methylation datasets from eight Japanese patients with CREBBP-related Rubinstein–Taybi syndrome (RSTS1_1–RSTS1_8) were included for comparison (25). DNA methylation data were processed according to the previously described analytical pipeline (25). In brief, methylated and unmethylated signal intensities were normalized with background and dye-bias correction, after which methylation levels at each CpG site were calculated as β-values (25).

Episignature analysis was performed using the previously reported MKHK-ID4-specific methylation probe set (19,20). Hierarchical clustering analysis was conducted using the Manhattan distance metric and the average linkage method based on probe-wise β values (25).

### Plasmid construction

A plasmid containing full-length human CREBBP cDNA cloned into the HaloTag®-N vector (Promega, Madison, WI, United States; FHC11778) was utilized in the analyses. R1868W, C1723fs, E1724D, and E1278K variants were generated through site-directed mutagenesis using the PrimeSTAR Mutagenesis Basal Kit (#R046A; Takara Bio). Primer sequences are summarized in Table S2.

### Immunoprecipitation assays for CREBBP acetyltransferase activity

To evaluate CREBBP acetyltransferase activity, immunoprecipitation assays were performed using modifications based on a method reported previously (25). HaloTag-fused wild-type CREBBP, R1868W, C1723fs, E1724D, E1278K, or mock constructs were transiently transfected into HEK293 cells using Lipofectamine 3000 (Thermo Fisher Scientific). Around 48 h after transfection, cells were harvested and lysed in RIPA buffer (#16488, Nacalai Tesque) with cOmplete™ Protease Inhibitor Cocktail (Roche Diagnostics). Lysates were incubated on ice for 30 min and then centrifuged at 12,000 rpm for 15 min.

To evaluate CREBBP autoacetylation, approximately 500 µg of total protein was incubated overnight at 4°C with CBP (D6C5) Rabbit Monoclonal Antibody Cell Signaling Technology; #7389, 1:2,000) with rotation, followed by incubation with Protein A Sepharose beads for 2 h. To evaluate acetylation of nonhistone proteins, lysates were incubated overnight at 4°C with Acetyl-Lysine Affinity Beads (Cat# AAC04-Beads, Cytoskeleton, Denver, CO, United States). After immunoprecipitation, the beads were rinsed three times with RIPA buffer and eluted by heating in 2× SDS sample buffer for 7 min.

Samples were separated on 4%–15% Mini-PROTEAN® TGX™ gels (Bio-Rad) at 120 V for 60 min and transferred to PVDF membranes at 100 V for 60 min.

For autoacetylation analysis, membranes were probed with CBP (D6C5) Rabbit Monoclonal Antibody (#7389, Cell Signaling Technology; 1:2,000) and anti-acetyl-lysine antibody [RM101]; (ab190479, Abcam, Cambridge, United Kingdom; 1:1,000). For nonhistone acetylation analysis, anti-acetyl-lysine antibody [RM101] alone was used. HRP-conjugated anti-rabbit IgG secondary antibody (#7074, Cell Signaling Technology; 1:2,000) was incubated for 90 min at the room temperature.

For input samples, CBP (D6C5) (#7389, 1:2,000) and GAPDH Rabbit Monoclonal Antibody (#2118, 1:2,000) were used.

### Histone extraction and Western blotting

Wild-type CREBBP, R1868W, C1723fs, E1724D, E1278K, or mock constructs were transiently transfected into HEK293 cells using Lipofectamine 3000 (Thermo Fisher Scientific). After 48 h, histone proteins were extracted using the Histone Extraction Kit (ab113476, Abcam). Samples were then separated on 4%–15% Mini-PROTEAN® TGX™ gels (Bio-Rad) at 120 V for 60 min and transferred to PVDF membranes at 100 V for 30 min.

Membranes were incubated overnight at 4°C with Histone H3 Rabbit Monoclonal Antibody (1:2,000; #4499 D1H2, Cell Signaling Technology) or Acetyl-Histone H3 (Lys27) Rabbit Monoclonal Antibody (1:2,000; #8173 D5E4, Cell Signaling Technology). HRP-conjugated anti-rabbit IgG secondary antibody (#7074, Cell Signaling Technology; 1:2,000) was incubated for 90 min at room temperature.

### Fluorescence recovery after photobleaching

COS7 cells transiently expressing HaloTag-fused CREBBP constructs (WT, R1868W, and C1723fs) were cultured on 35-mm glass-bottom dishes (Matsunami Glass, Osaka, Japan). After 24 h, cells were labeled with HaloTag TMR ligand (1:200 dilution; Promega, Madison, WI, United States). Prior to imaging, the culture medium was replaced with FluoroBrite DMEM (Thermo Fisher Scientific).

FRAP analysis was performed on a confocal laser-scanning microscope (FV3000, Olympus, Tokyo, Japan) under 37°C and 5% CO₂ conditions.

Given that wild-type CREBBP condensates were relatively small, full FRAP was conducted in regions approximately 1.4 µm in diameter, in which the entire condensate was photobleached. This approach enabled the evaluation of the molecular exchange between condensates and the surrounding nucleoplasm rather than the diffusion within condensates.

For R1868W experiments, cells were treated with either DMSO or A485 (3 µM; Selleck Chemicals, Houston, TX, United States) for 1 h prior to FRAP analysis. Fluorescence recovery curves were normalized to pre-bleach intensity and corrected for background fluorescence and acquisition bleaching.

Mobile fraction and half-time of recovery (τ1/2) derived from FRAP curves were compared between the groups using the Mann–Whitney U test. For comparisons among WT, R1868W, and C1723fs, P values were adjusted for multiple pairwise comparisons using the Holm method separately for each parameter. A p value of <0.05 indicated statistical significance. Graphs were annotated as follows: *p < 0.05, **p < 0.01, ***p < 0.001, and ns (not significant).

### Time-lapse imaging

COS7 cells transiently expressing HaloTag-fused CREBBP R1868W were treated with either DMSO or A485 for 1 h following 24 h of expression and subjected to live-cell time-lapse imaging to evaluate nuclear condensate dynamics. Imaging was performed using an FV3000 confocal laser-scanning microscope (Olympus) equipped with a 100× oil-immersion objective lens. Cells were maintained at 37°C and 5% CO₂ during imaging. To compensate for focal drift during long-term imaging, Z drift compensation (ZDC) was applied every 15 s. Time-lapse images were acquired in a single focal plane at intervals of 10–30 s for up to 80–160 min.

Condensate fusion events and formation of dumbbell-shaped intermediates were assessed visually.

Different treatment conditions were not imaged simultaneously, and each condition was analyzed independently. Moreover, given that cell movement occasionally limited long-term tracking of individual condensates, quantitative analyses (e.g., aspect ratio measurements) were not performed. Therefore, this analysis was used primarily for qualitative evaluation of condensate dynamics.

### Immunocytochemistry and BRD4 recruitment analysis

COS7 cells transiently expressing HaloTag-fused CREBBP R1868W were labeled with HaloTag TMR ligand (1:200 dilution; Promega) 24 h after transfection. Following treatment with either DMSO or JQ1 (1 µM for 1 h; Selleck Chemicals), cells were fixed with 4% paraformaldehyde for 10 min, permeabilized with 1% Triton X-100, and then blocked with 10% goat serum for 1 h. Cells were cultivated overnight at 4 °C with anti-BRD4 antibody (1:500; Abcam, Cambridge, United Kingdom; ab128874). Alexa Fluor 488-conjugated secondary antibody (1:500; Thermo Fisher Scientific) was used for detection.

BRD4 localization around condensates was evaluated using radial fluorescence intensity distribution analysis. Distances from the condensate center were normalized to the condensate radius, whereas BRD4 fluorescence intensities at each distance were normalized to the mean nuclear fluorescence intensity.

For H3K27ac immunostaining, COS7 cells transiently expressing HaloTag-fused CREBBP R1868W were treated with DMSO and subjected to the same fixation, permeabilization, and blocking procedures. Cells were incubated for 1 h at room temperature with mouse monoclonal anti-H3K27ac antibody (1:500; Active Motif) and rabbit polyclonal anti-HaloTag antibody (1:500; Promega). Species-specific fluorescent secondary antibodies were used for detection.

## Data availability

Data supporting the experimental and quantitative analyses, including uncropped and unedited Western blot images, are included in the article and/or Supporting Information. Individual-level whole-exome sequencing and genome-wide DNA methylation data are not publicly available, because their secondary use and sharing are not covered by the current participant consent. Raw microscopy images underlying the quantitative analyses and the code used for data analysis are available from the corresponding author upon reasonable request.

## Acknowledgements

We would like to express sincere thanks to the patient families and medical staff. We would also like to thank Ms. Nana Kobayashi and Yukimi Abe for sample preparation and data processing assistance. This study was supported in part by grants from the Initiative on Rare and Undiagnosed Diseases (IRUD) of the Japan Agency for Medical Research and Development (AMED) (JP22ek0109549, JP24ek0109760, JP25ek0109760, JP26ek0109760, JP26ek0109815 and JP26ek0109617 to T.Kan.) and by a grant from the National Center for Child Health and Development (Grant No. 2024B-18 to T.Kan.).

## Notes

### Competing Interest Statement

The authors have declared no competing interest.

### Author Declarations

This study was approved by the Ethics Committee of the National Center for Child Health and Development (Approval No. 2020-326).

## References

1. C.P. Brangwynne, et al., Germline P granules are liquid droplets that localize by controlled dissolution/condensation. Science 324, 1729–1732, (2009).

2. M. Feric M, et al., Coexisting liquid phases underlie nucleolar subcompartments. Cell 165, 1686–1697, (2016).

3. B.R. Sabari, et al., Coactivator condensation at super-enhancers links phase separation and gene control. Science 361, eaar3958, (2018).

4. Y.E. Guo, et al., Pol II phosphorylation regulates a switch between transcriptional and splicing condensates. Nature 572, 543–548, (2019).

5. P.J. Flory, Thermodynamics of high polymer solutions. J Chem Phys 10, 51–61, (1942).

6. Y. Shin, C.P. Brangwynne, Liquid phase condensation in cell physiology and disease. Science 357, eaaf4382, (2017).

7. J. Wang, et al., A molecular grammar governing the driving forces for phase separation of prion-like RNA binding proteins. Cell 174, 688–699, (2018).

8. K.M. Ruff, et al., Molecular grammars of predicted intrinsically disordered regions that span the human proteome. Cell 189, 323–342, (2026).

9. H.R. Kilgore, et al., Protein codes promote selective subcellular compartmentalization. Science 387,1095–1101, (2025).

10. S.F. Banani, et al., Genetic variation associated with condensate dysregulation in disease. Dev Cell 57, 1776–1788, (2022).

11. C.H. Li, et al., MeCP2 links heterochromatin condensates and neurodevelopmental disease. Nature 586, 440–444, (2020).

12. M.A. Mensah, et al., Aberrant phase separation and nucleolar dysfunction in rare genetic diseases. Nature 614, 564–571, (2023).

13. E. Ortega, et al., Transcription factor dimerization activates the p300 acetyltransferase. Nature 562, 538–544, (2018).

14. L. Ma, et al., Co-condensation between transcription factor and coactivator p300 modulates transcriptional bursting kinetics. Mol Cell 81, 1682–1697, (2021).

15. Z. Ibrahim, et al., Structural insights into p300 regulation and acetylation-dependent genome organisation. Nat Commun 13, 7759, (2022).

16. D. Yu, et al., Structural mechanism of BRD4-NUT and p300 bipartite interaction in propagating aberrant gene transcription in chromatin in NUT carcinoma. Nat Commun 14, 378, (2023).

17. F. Petrij, et al., Rubinstein-Taybi syndrome caused by mutations in the transcriptional co-activator CBP. Nature 376, 348–351, (1995).

18. L.A. Menke, et al., CREBBP mutations in individuals without Rubinstein-Taybi syndrome phenotype. Am J Med Genet A, 170, 2681–2693, (2016).

19. S. Haghshenas, et al., Menke-Hennekam syndrome: Delineation of domain-specific subtypes with distinct clinical and DNA methylation profiles. HGG Adv 5, 100287, (2024).

20. M.A. Levy, et al., Novel diagnostic DNA methylation episignatures expand and refine the epigenetic landscapes of Mendelian disorders. HGG Adv 3,100075, (2021).

21. A. Piai, et al., Just a flexible linker? The structural and dynamic properties of CBP-ID4 revealed by NMR spectroscopy. Biophys J, 110, 372–381, (2016).

22. B. Xue, et al., PONDR-FIT: A meta-predictor of intrinsically disordered amino acids. Biochim Biophys Acta 1804, 996–1010, (2010).

23. K.L. Gelder, et al., CBP-IDRs regulate acetylation and gene expression. Cell Rep 45, 117109, (2026).

24. E. Kalkhoven, et al., Loss of CBP acetyltransferase activity by PHD finger mutations in Rubinstein-Taybi syndrome. Hum Mol Genet 12, 441–450, (2003).

25. T. Kawai, et al., DNA methylation data from Japanese patients with Rubinstein–Taybi syndrome. Hum Genome Var 12, 32, (2025).

26. T. Aoki, et al., Functional analysis of novel and recurrent *RINT1* variants in patients with infantile liver dysfunction. J Hum Genet 71, 23–34, (2026).

27. M. Kumar, et al., Inhibition of histone acetyltransferase function radiosensitizes CREBBP/EP300 mutants via repression of homologous recombination, potentially targeting a gain of function. Nat Commun 12, 6340, (2021).

28. E. Nishi, et al., The novel and recurrent variants in exon 31 of CREBBP in Japanese patients with Menke-Hennekam syndrome. Am J Med Genet A 188, 446–453, (2022).

