## Supporting information 1 for "*CREBBP* variants underlying Menke–Hennekam syndrome exhibit histone acetyltransferase hyperactivity with divergent phase behaviors"

**This PDF file includes:**

Fig.S1 to S6

Table S1 to S2

SI References

Supplementary Figures

Fig. S1. Clinical features and pedigrees of the two families with MKHK-associated CREBBP variants.

Fig. S2. Splicing and protein expression analyses of the novel CREBBP exon 30 terminal variant identified in F2.

Fig. S3. DNA methylation episignature analysis using MKHK-ID4–specific probes.

Supplementary Tables

Fig. S4. Peri-condensate accumulation of H3K27ac around CREBBP R1868W condensates

Fig. S5. Uncropped Western blot images corresponding to Figure 1C and D.

Fig. S6. Uncropped Western blot images corresponding to Figure 1E.

Table S1. Detailed information on CREBBP variants identified in individuals F1 and F2.

Table S2. Primer sequences used for cDNA sequencing and site-directed mutagenesis.

SI References

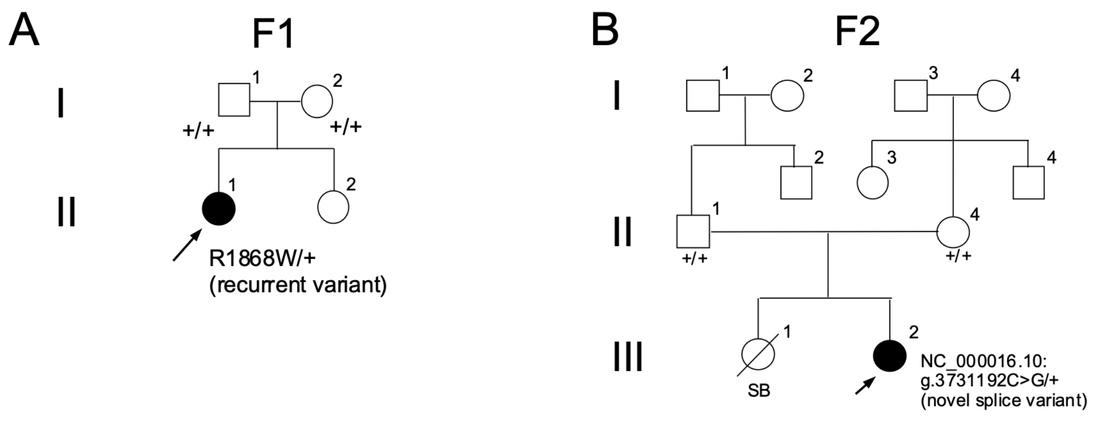


Fig.S1

**Fig. S1 Pedigrees of the two families with MKHK-associated CREBBP variants**

(A) Pedigree of Family 1 (F1) carrying the recurrent missense variant (*CREBBP*: NM_004380: c.5602C>T, p.(Arg1868Trp).

(B) Pedigree of Family 2 (F2) carrying the novel CREBBP splice variant (NC_000016.10:g.3731192C>G).

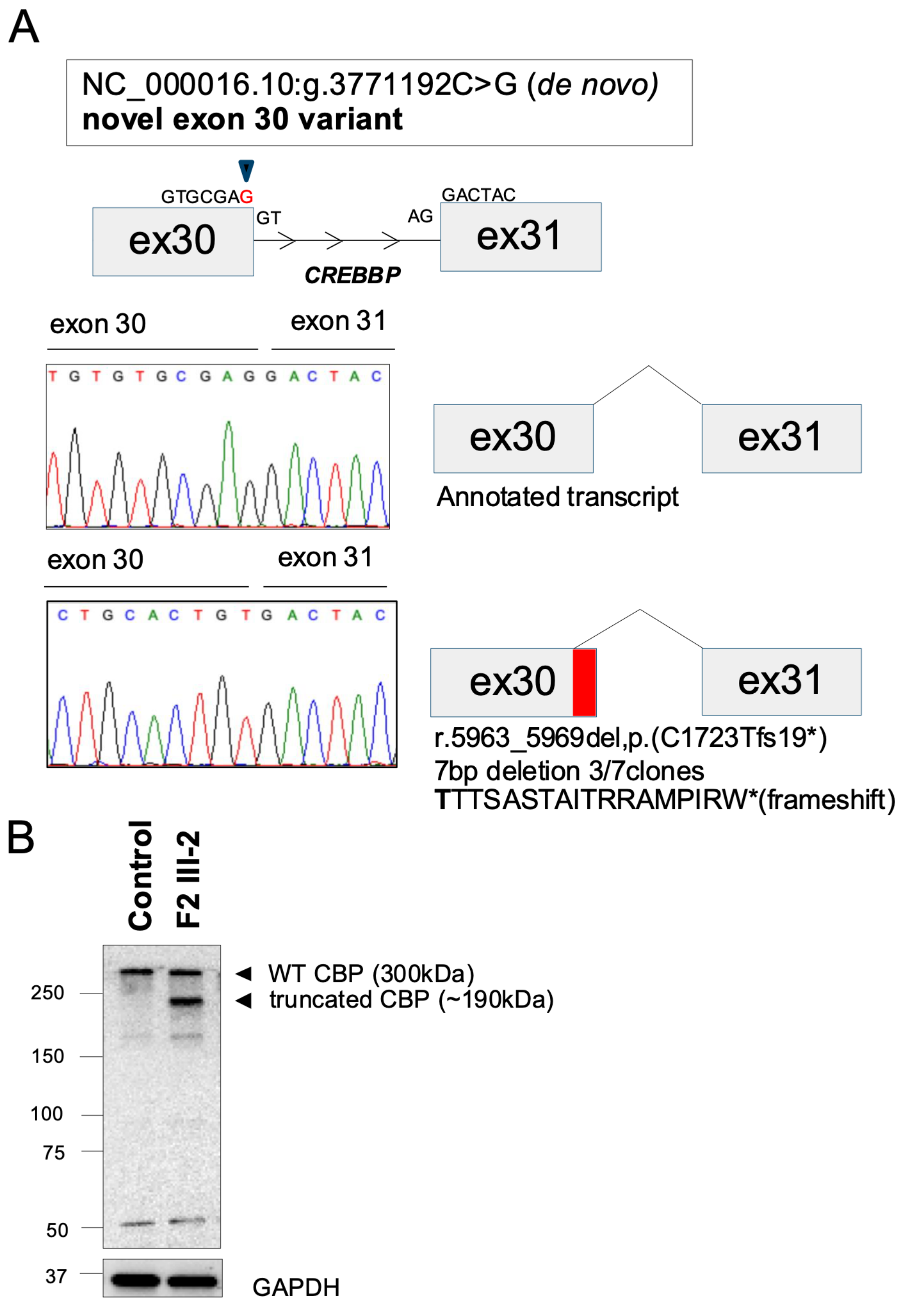


Fig.S2

**Fig. S2 Splicing and protein expression analyses of the novel *CREBBP* exon 30 terminal variant identified in F2**

(A) cDNA sequencing and TA cloning analysis of the *CREBBP* exon 30 terminal variant, identified in the F2 proband (III-2). cDNA was generated from peripheral blood RNA, followed by PCR amplification spanning exons 29–31 and TA cloning analysis. Sequencing of seven independent clones identified two aberrant transcript species. Three of seven clones contained a 7-bp deletion resulting in a frameshift transcript (*CREBBP*: NM_004380.3:r.5963_5969del, p.(Cys1723Thrfs*19) [C1723fs]), whereas one of seven clones retained normal splicing but carried a single nucleotide substitution transcript (*CREBBP:* NM_004380.3:r.5969g>c, p.(Glu1724Asp) [E1724D]). The remaining clones corresponded to wild-type transcripts.

(B) Western blot analysis of CREBBP protein expression in lymphoblastoid cell lines (LCLs), derived from the F2 proband (III-2) and a healthy control. Aside from the full-length CREBBP (~300 kDa), a truncated CREBBP protein of approximately 190 kDa was detected in the F2 sample. GAPDH was employed as a loading control. These findings suggest that the C1723fs transcript escapes nonsense-mediated mRNA decay and is expressed as a truncated protein product.

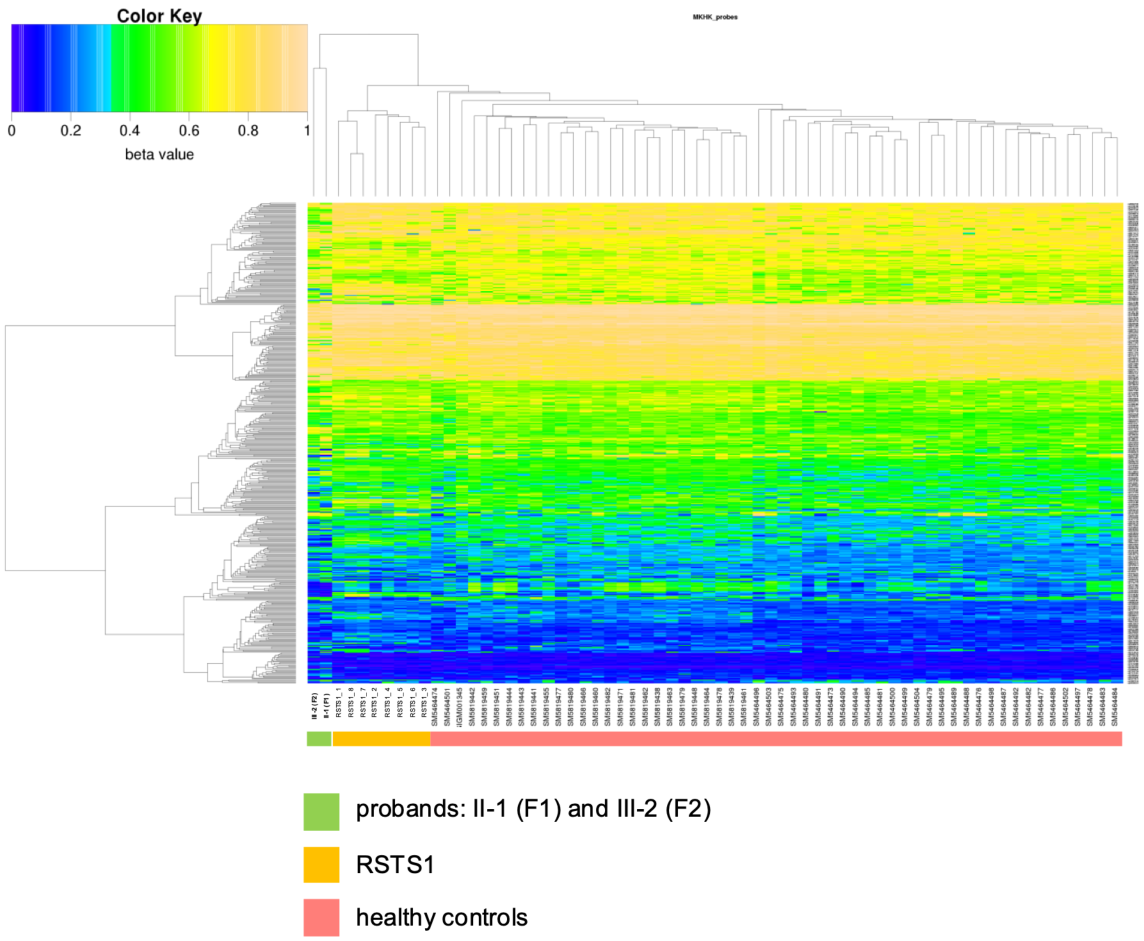


Fig.S3

**Fig. S3 DNA methylation episignature analysis using MKHK-ID4–specific probes**

Hierarchical clustering of probe-wise DNA methylation β values from the two MKHK probands (F1 II-1 and F2 III-2), eight Japanese patients with **CREBBP-related Rubinstein–Taybi syndrome** (RSTS1_1–RSTS1_8) reported previously (1), and healthy controls. Healthy controls consisted of two in-house controls (one adult female patient and one female child) together with 55 publicly available healthy pediatric controls from GSE180534 and GSE193730. Clustering was performed using the Manhattan distance metric and the average linkage method. Heatmap colors represent DNA methylation β values ranging from 0 (blue) to 1 (yellow).

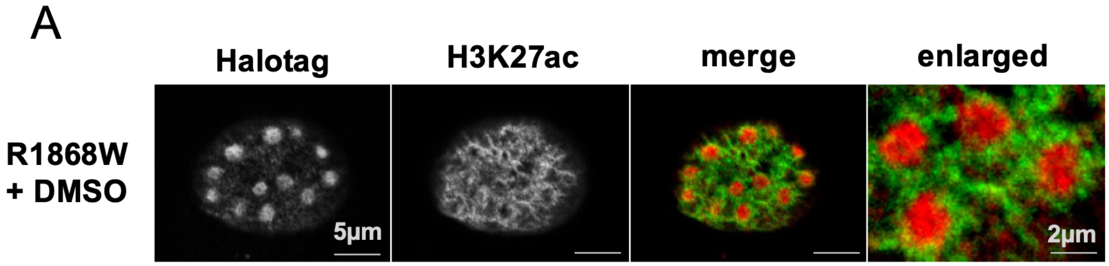


Fig.S4

**Figure S4. Peri-condensate accumulation of H3K27ac around CREBBP R1868W condensates**

Representative single optical sections of COS7 cells transiently expressing HaloTag-fused CREBBP R1868W and immunostained for H3K27ac. H3K27ac signals were observed predominantly in regions surrounding R1868W condensates. From left to right: HaloTag, H3K27ac, and merged images (scale bar, 5 μm). The rightmost panel shows an enlarged merged image of condensates (scale bar, 2 μm).

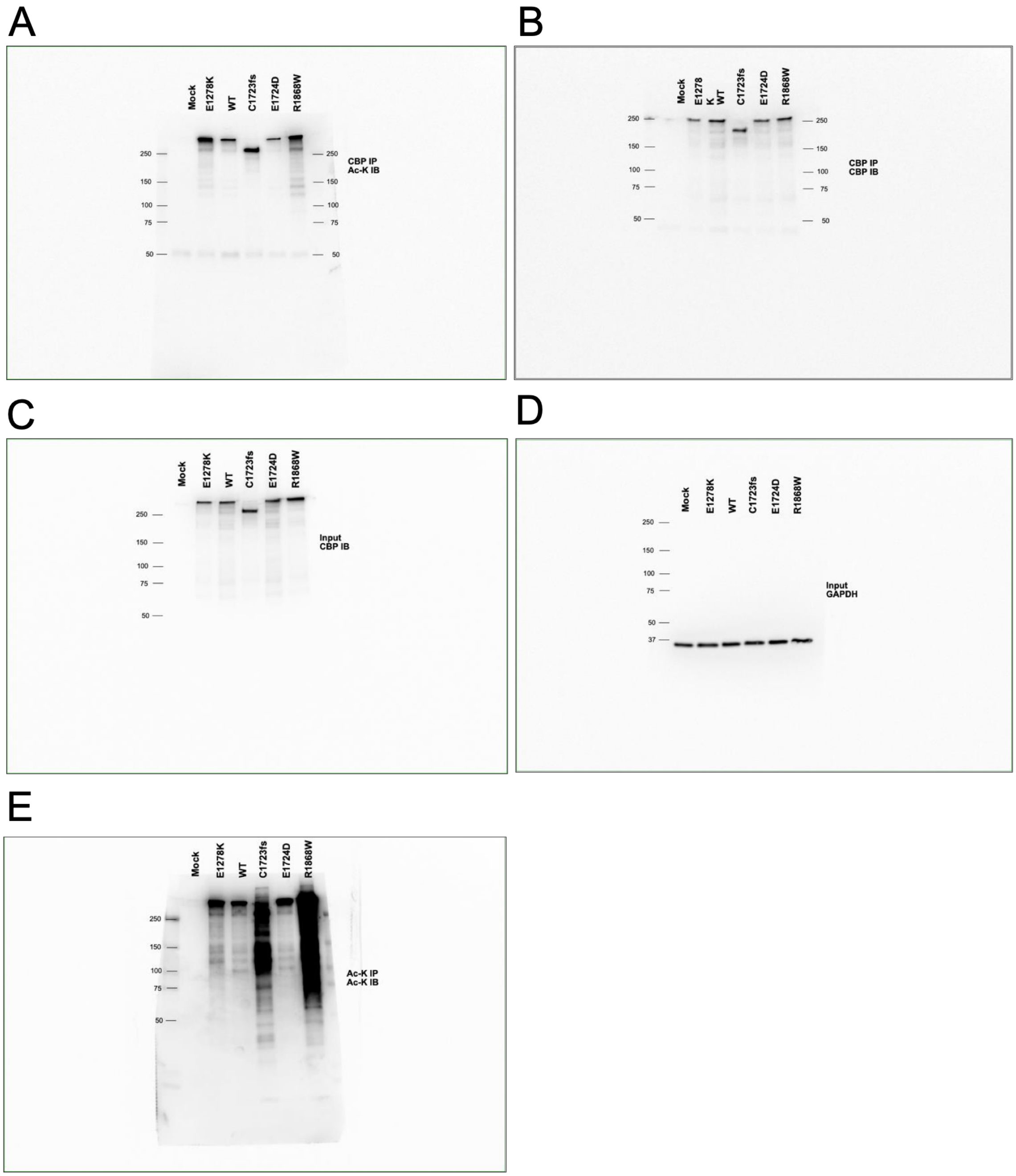


Fig.S5

**Fig. S5. Uncropped Western blot images corresponding to Figures 1C and D**

(A) CBP IP, Ac-K IB. (B) CBP IP, CBP IB. (C) Input, CBP IB. (D) Input, GAPDH. (E) Ac-K IP, Ac-K IB. Molecular weight labels were assigned according to the migration of the dual-color protein ladder run on the same gel.

A

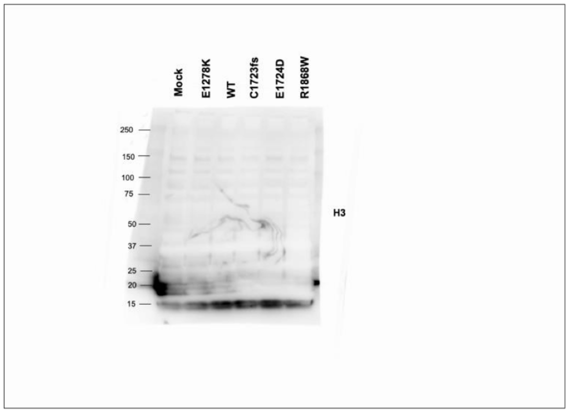


B

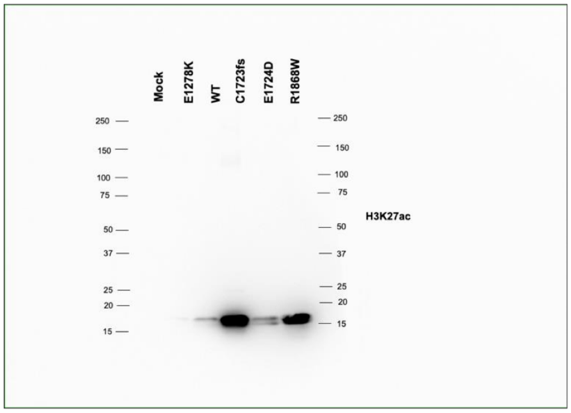


Fig.S6

**Fig. S6. Uncropped Western blot images corresponding to Figure 1E**

(A) H3. (B) H3K27ac. Molecular weight labels were assigned according to the migration of the dual-color protein ladder run on the same gel.

### Tables

**Table S1. Detailed information on CREBBP variants identified in individuals F1 and F2.**

| Patient | Gene | Chr | POS | REF | ALT | Reference Sequence | Nucleotide | Protein | Inheritance |
|---|---|---|---|---|---|---|---|---|---|
| F1 (II-1) | *CREBBP* | 16 | 3729445 | G | A | NM_004380.3 | c.5602C>T | p.(R1868W) | *de novo* |
| F2(III-2) | *CREBBP* | 16 | 3731192 | C | G | NM_004380.3 | r.[5963_5969del,5969g>c] | p.(C1723Tfs19*,E1724D) | *de novo* |

| Patient | SIFT | Polyphen-2 | CADD | AlphaMissense | In-house | gnomAD v4.1.0 |
|---|---|---|---|---|---|---|
| F1 (II-1) | Deleterious | probably damaging | 25 | Likely Pathogenic | 0 | NA |
| F2(III-2) | NA/Tolerated | NA/probably damaging | 34 | NA/Likely Pathogenic | 0 | NA |

| Patient | dbSNP | ToMMo 54KJPN | ClinVar Clinical Significance | ACMG evidence criteria | Classification |
|---|---|---|---|---|---|
| F1 (II-1) | rs886039491 | NA | Pathogenic | PS2,PS3,PM2,PP4 | Pathogenic |
| F2(III-2) | 0 | NA | NA | PS2, PS3,PM2,PM4 | Pathogenic |

*CREBBP* variants identified in individuals F1 (II-1) and F2 (III-2) are listed with genomic coordinates, reference sequence, nucleotide and protein changes, inheritance, in silico pathogenicity predictions, allele frequencies in population databases, dbSNP identifiers, ClinVar classification, ACMG evidence criteria, and final variant classification. Allele frequencies were assessed using an in-house whole-exome sequencing database of 2,714 individuals at the National Center for Child Health and Development, gnomAD v4.1.0, and the ToMMo 54KJPN database. In silico predictions included SIFT, PolyPhen-2, CADD, and AlphaMissense, where applicable. ACMG evidence criteria and variant classifications were assigned based on population frequency data, public variant databases, inheritance, clinical findings, and functional analyses performed in this study. NA, not available or not applicable; CADD, Combined Annotation Dependent Depletion.

**Table S2. Primer sequences used for cDNA sequencing and site-directed mutagenesis**

| Primer ID | Sequence |
|---|---|
| **cDNA sequencing primer** | |
| CREBBP_ex29_F | ACGTGTCCAATGACCTGTCC |
| CREBBP_ex31_R | GCCCCCACTTCACCATCTTA |
| **Cloning primer** | |
| CREBBP_R1868W_mut_F | TGCGCCGGTGGATGGCCACCATGAACACC |
| CREBBP_R1868W_mut_R | CCATCCACCGGCGCATGAGCTGGGCCTG |
| CREBBP_C1723Tfs19*_mut_F | GCACTGTGACTACGACCTCTGCATCAACT |
| CREBBP_C1723Tfs19*_mut_R | TCGTAGTCACAGTGCAGTGCCAGCG |
| CREBBP_E1724D_mut_F | TGTGCGACGACTACGACCTCTGCATCAACT |
| CREBBP_E1724D_mut_R | CGTAGTCGTCGCACACAGTGCAGTGCCAG |
| CREBBP_E1278K_mut_F | AGACCCCAAACCTTTCGTTGATTGCAAG |
| CREBBP_E1278K_mut_R | AAAGGTTTGGGGTCTAAGGTATCATTTTTCT |

Primer sequences used for cDNA sequencing and site-directed mutagenesis are summarized together with their respective applications. cDNA sequencing primers were used to amplify and sequence the CREBBP transcript region spanning exons 29–31. Mutagenesis primers were used to generate CREBBP constructs harboring R1868W, C1723Tfs*19, E1724D, or E1278K variants. All primer sequences are presented in the 5′ to 3′ direction.

### SI References


1. T. Kawai, et al. DNA methylation data from Japanese patients with Rubinstein–Taybi syndrome. *Hum Genome Var*, 12(1), 27, (2025).
